# Autism spectrum disorder common variants associated with regional lobe volume variations at birth: cross-sectional study in 273 European term neonates in developing Human Connectome Project

**DOI:** 10.1101/2024.08.18.24311983

**Authors:** Hai Le, Daphna Fenchel, Konstantina Dimitrakopoulou, Hamel Patel, Charles Curtis, Lucilio Cordero-Grande, A. David Edwards, Joseph Hajnal, Jacques-Donald Tournier, Maria Deprez, Harriet Cullen

**Affiliations:** Centre for the Developing Brain, Perinatal Imaging and Health Department, King’s College London, London, United Kingdom; Translational Bioinformatics Platform, NIHR Biomedical Research Centre Guy’s and St. Thomas’ NHS Foundation Trust and King’s College London, London, United Kingdom; NIHR BioResource Centre Maudsley, NIHR Maudsley Biomedical Research Centre at South London and Maudsley NHS Foundation Trust & Institute of Psychiatry, Psychology and Neuroscience, King’s College London, London, United Kingdom; Social Genetic & Developmental Psychiatry Centre, Institute of Psychiatry, Psychology and Neuroscience, King’s College London, London, United Kingdom; Biomedical Image Technologies, ETSI Telecomunicación, Universidad Politécnica de Madrid & CIBER-BBN, ISCIII, Madrid, Spain; Department of Medical and Molecular Genetics, King’s College London, London, United Kingdom

## Abstract

Increasing lines of evidence suggest cerebral overgrowth in autism spectrum disorder (ASD) children in early life, but few studies have examined the effect of ASD common genetic variants on brain volumes in a general paediatric population. This study examined the association between ASD polygenic risk score (PRS) and volumes of the frontal, temporal, parietal, occipital, fronto-temporal and parieto-occipital lobes in 273 term-born infants of European ancestry in the developing Human Connectome Project. ASD PRS was positively associated with frontal (β = 0.027, p_FDR_ = 0.04) and fronto-temporal (β = 0.024, p_FDR_ = 0.01) volumes, but negatively with parietal (β = −0.037, p_FDR_ = 0.04) and parieto-occipital (β = −0.033, p_FDR_ = 0.01) volumes. This preliminary result suggests potential involvement of ASD common genetic variants in early structural variations linked to ASD.

## 1. Introduction

Early neuroanatomical differences have been associated with autism spectrum disorder (ASD) ^1^, but the role of ASD common genetic variants on normal brain development in neonates remains unclear. Preliminary examination of additive effects of the single nucleotide polymorphisms (SNPs) as measured by polygenic risk scores (PRS) has suggested links between ASD polygenic risk and structural abnormalities in young children. Recently, in two general paediatric populations between 9-11 and 3-14 years of age, findings revealed associations between ASD PRS and regional cortical thickness ^2^ and gyrification ^3^. Despite small effect sizes, these results demonstrate possible brain morphological alterations associated with ASD common genetic variants.

Increasing lines of evidence suggest early brain structural variations are likely to precede ASD diagnosis and can be detected in the first two years of life ^4^. For instance, greater extra-axial cerebral spinal fluid ^5^, hyper-expansion of cortical surface area ^1^, and ventriculomegaly ^6^ can be found in infants with ASD or at high familial risk of the disease as early as six months after birth. Similarly, total brain tissue and regional lobe volumes appear larger in ASD cases between two to four years old as compared with controls ^7–9^. Further evidence suggests these regional lobe volume variations may not be consistent throughout the cerebrum and that frontal and temporal lobes are relatively more affected than parietal and occipital lobes ^10–13^. Taken together, this points toward possible disruption to the normal neurodevelopmental process (i.e., from primary to more complex functions) in the disease ^14^.

Common genetic variants appear to explain at least 12% of the heritability of ASD ^15^. Functional annotation of the genetic loci revealed an enrichment in genes particularly important during fetal corticogenesis ^15,16^. Similarly, recent transcriptomic analysis of postmortem ASD tissues spanning across four cortical lobes found evidence of variable changes in genetic expression of ASD risk genes along the anterior-posterior axis ^17^ reflecting possible alterations to fundamental elements of cortical organisation in the disease. Concurrently, the neonatal brain is most rapidly changing immediately after birth ^18,19^. The first two postnatal years are characterised by dynamic, regionally non-uniform and protracted brain growth ^18^. Disruption to normal development during this critical window as a result of either genetic or environmental factors may have long lasting effects on brain structure and function ^20^.

Given the evidence of regional lobe volume variations associated with the disease and possible involvement of ASD risk genes in early neurodevelopmental process ^15,17^, the current work aimed to the examine the association between ASD common variants and regional lobe volumes in the general population at birth.

## 2. Methods

### 2.1. Cohort

The data used were analysed from infants, who were recruited from St. Thomas’ Hospital London, UK as part of the third release of developing Human Connectome Project (dHCP) (https://biomedia.github.io/dHCP-release-notes/) ^21^. The dHCP was conducted according to the principles of the Declaration of Helsinki and ethical approval was given from the UK National Research Ethics Service. Written parental consent was provided for all subjects.

#### 2.1.1 Genotyping and genetic quality control

The genetic data quality control and preprocessing steps used in this study are described in ^22^. Briefly, of the 842 saliva samples, genotype data were collected (Oragene DNA OG-250 kit) and genotyped for SNPs genome-wide on the Illumina Infinitum Omni5-4 v1.2 array. If multiple samples were provided, only one per individual was retained for analysis (randomly chosen). Excluded were also genotyped data based on the following criteria: completeness less than 95%, gender discrepancy and genotyping failure of more than 1% of the SNPs. If the relatedness score between any individual pair was above a cut-off (pi_hat >= 0.1875), only one sample was randomly retained. Finally, SNPs were filtered based on the following criteria: being non-autosomal, having minor allele frequency less than 0.05, missing in more than 1% of individuals or deviating from Hardy-Weinberg equilibrium with a p-value < 1 x 10^-5^. This resulted in a sample of 754 individuals with high-quality genetic data.

### 2.2. Data pre-processing

Given the strong effect of prematurity on the imaging phenotype, out of 887 available imaging scans, only those of term-born infants (born after at least 37 weeks of gestational age (GA)) were selected for analysis (n=582 scans). Briefly, infants brain scans were obtained during their natural sleep using a dedicated neonatal brain imaging system on a 3T Philips Achieva scanner ^23^. Here, T2-weighted MRI were acquired using a TSE sequence with parameters TR = 12 s, TE = 156 ms, and resolution (mm) 0.8 x 0.8 x 1.6. A series of motion correction ^24^ and super-resolution reconstruction ^25^ techniques were then employed to produce images of resolution (mm) 0.5 x 0.5 x 0.5. Subsequently, the T2 images were segmented with the DrawEM neonatal segmentation algorithm (https://github.com/MIRTK/DrawEM) ^26^. This technique utilized spatial priors of 50 brain regions in the form of 20 manually segmented atlases ^27^ in combination with tissue segmentation using an Expectation-Maximisation technique to model intensities of different tissues classes and their subdivisions. This allowed the images to be accurately parcellated into 87 regions (Figure 1A). The quality control was performed as a part of dHCP minimal processing pipeline ^28^. The absolute volumes of each structure were calculated as the total number of voxels multiplied by the voxel dimension ^29^. Images with radiology score of 5 (i.e, major brain lesions) denoted by clinical experts were excluded (26 scans removed). Finally, of the duplicate scans, the first imaging session was retained (2 scans removed).

**Figure 1.**
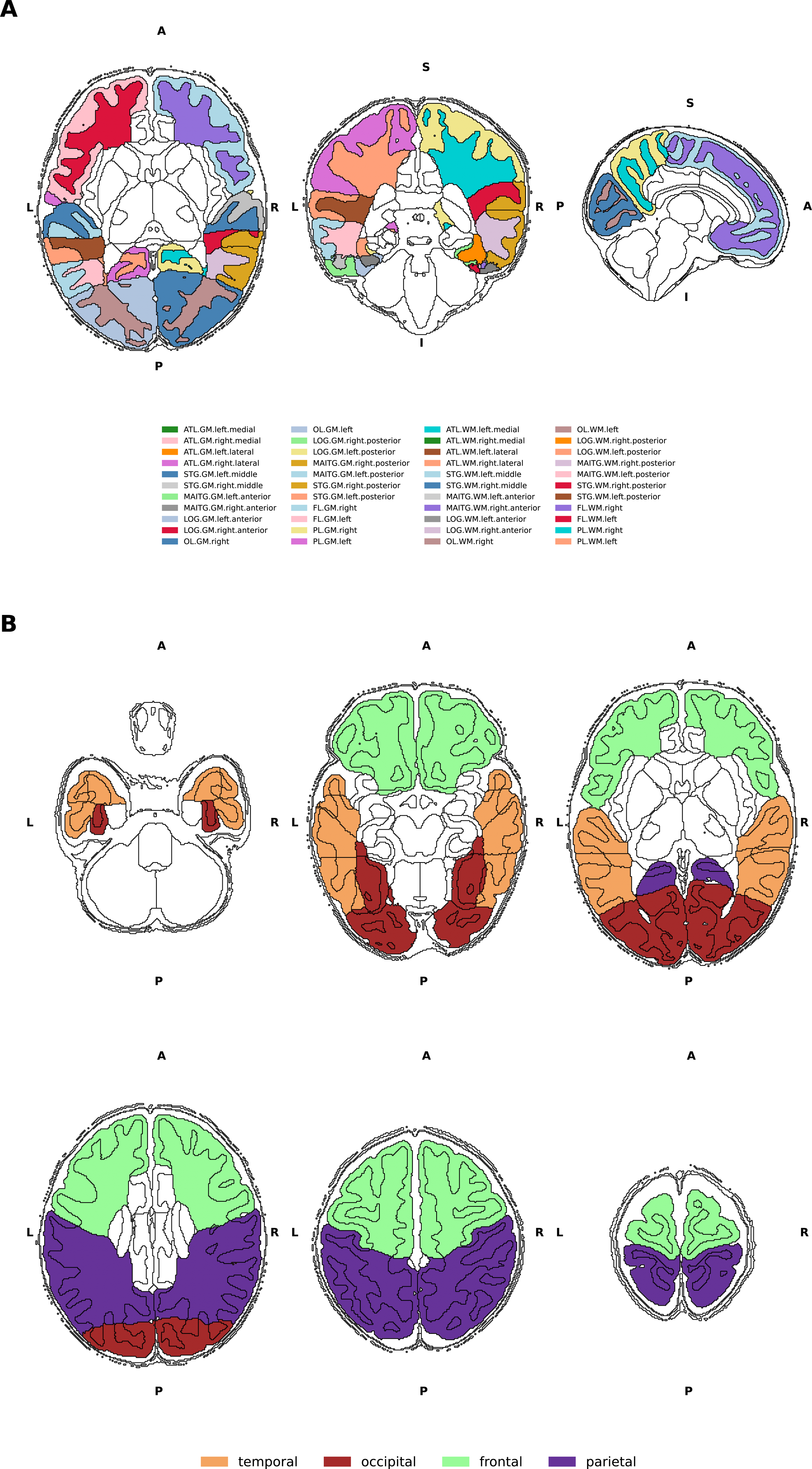
Visualisation of DrawEM parcellations. A-Visualisation of the brain regions examined in axial, coronal and sagittal views. Black outline denotes segmented brain regions. Regions found within the four cortical lobes examined in this study are coloured. B-Six axial views from inferior to posterior indicating the boundaries of the four cortical lobes. ATL – anterior temporal lobe, GPEA-gyri paraphippocampalis et ambiens, STG-superior temporal gyrus, MAITG – medial and inferior temporal gyrus, LOGGF – lateral occipotemporal gyri, gyrus fusiform, CG-cingulate gyrus, FL – frontal lobe, PL – parietal lobe, OL – Occipital lobe, INSU-Insula. WM – white matter, GM – grey matter. A – Anterior, P – Posterior, S-Superior, I-Inferior, L – Left, R-Right.

#### 2.2.1 Volumetric data

From the original 87 segmented regions, the cortical grey and white matter regions that made up the frontal, temporal, occipital and parietal lobes were selected in accordance with previous studies ^13,30^. Here, the total grey and white matter volumes in both hemispheres of six brain regions – frontal, temporal, occipital, parietal, fronto-temporal and parieto-occipital were calculated. In addition, the brain volumes of the superior temporal gyrus (middle and posterior part), the inferior temporal gyri (anterior and posterior part) and the anterior temporal lobe (medial and lateral part) were summed up to designate the temporal lobe. The lateral occipitotemporal gyrus and gyrus fusiform (posterior and anterior part) were included in the volume of the occipital lobe (Figure 1B). The total brain tissue volume (TBV) was computed as the sum of the volumes of cortical white and grey matter, deep grey matter, cerebellum and brainstem.

#### 2.2.2 Populations stratification

Ancestry subpopulations were identified by merging our cohort of 754 infants with 2504 individuals from the 1000 Genomes Project ^31^ using a subset of common autosomal SNPs. Principal component analysis (PCA) was then performed on the resulting genetic dataset with PLINK and principal components (PCs) generated were plotted to visually assign individuals to ancestral subgroups. Since the discovery sample used in the genome-wide association study included exclusively Danish individuals, only infants of European ancestry were chosen for the ongoing analysis. Here, 429 (57%) infants were determined to have European ancestry.

European ancestry PCs were derived from the genetic data of the 429 infants using PCA, and visual examination of the PC pairwise scatterplots was carried out to exclude European ancestral group outliers (6 individuals excluded). Finally, of the remaining infants of European ancestry, 273 were born at term and examined in this study (Table 1; Figure 2).

**Figure 2.**
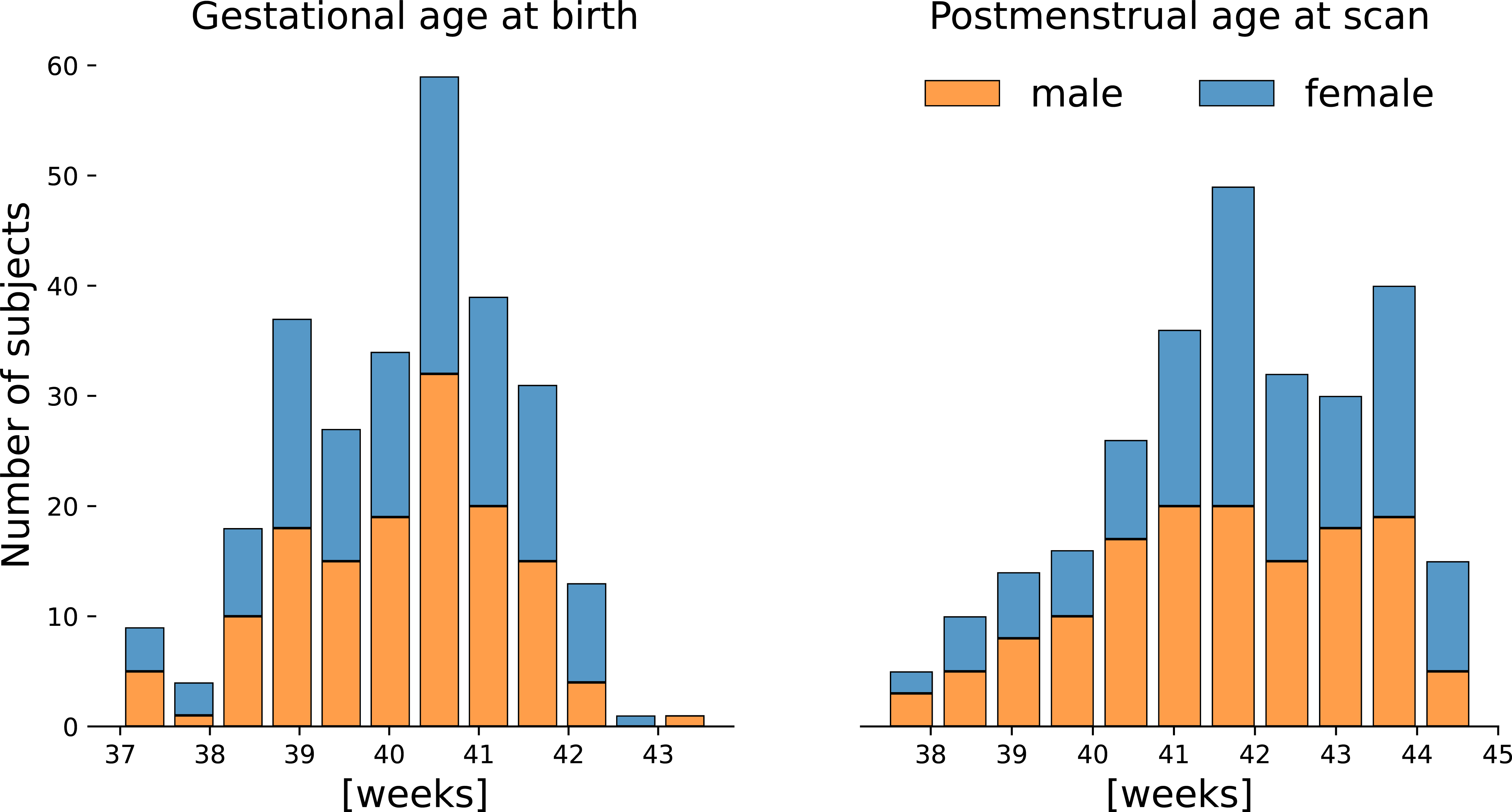
Term-born European neonatal cohort. Distribution of gestational age at birth and postmenstrual age at scan of participants included in the study.

**Table 1.** Demography of European neonatal cohort included in this study.

|  |  |
| --- | --- |
| Individuals | 273 |
| Female/Male | 133/140 |
| GA (weeks) | 40.16±1.22 |
| PMA (weeks) | 41.75±1.66 |
| Total Brain Volume (mm<sup>3</sup>) | 380251±49469 |
| Birth weight (kg) | 3.48±0.47 |

#### 2.2.3 Polygenic risk score

The summary statistics used to determine individual PRS were derived from the largest to date ASD GWAS ^15^. This analysis was performed on 18 381 individuals with ASD and 27 969 controls from a unique Danish population ^32^. PRS for each dHCP infant were estimated using PRSice-2 at 10 different p-value thresholds (P_T_): 10^-8^, 10^-6^, 10^-5^, 0.0001, 0.001, 0.01, 0.05, 0.1, 0.5, and 1, such that each score was composed of only those SNPs with ASD GWAS association p-value less than the respective threshold. Genotype data of European individuals in the 1000 Genomes project was used as the external linkage disequilibrium reference panel ^31^. PCA was then carried out on all 10 raw PRS values to obtain the first principal component (PRS-PC1) ^33^. In accordance with several genetic-neuroimaging studies, this unsupervised approach was selected due to its ease of implementation and reduction of multiple testing without loss of power ^34–36^. In the present study, PRS-PC1 explained 44% of the total variance in PRS scores and was positively associated with PRS at all P_T_ (Supplementary Figure 1).

### 2.3. Statistical Analysis

For each of the 6 dependent variables of interest (frontal, temporal, occipital, parietal, fronto-temporal, parieto-occipital lobe volumes), a linear regression was performed with PRS-PC1 as the independent variable, and the first 3 European ancestry PC, GA, postmenstrual age at scan (PMA), sex and TBV as covariates (i.e., Volume = PRS-PC1 + Ancestry + GA + PMA + Sex + TBV). The multiple-testing correction was carried out using false discovery rate (FDR) method. Results with p-value_FDR_< 0.05 were considered statistically significant.

Given our relatively small cohort, the results were further examined using three additional stability tests. Firstly, the sample was randomly divided into 2 data sets. PCA of the PRS and subsequent regression analysis described above were carried out on both data sets separately, the results of which were then compared. Here, the result was considered consistent if both sets yielded associations reported in the primary analysis. This test was repeated 1000 times using different random splits. Secondly, brain volume of the top and bottom 20% of the PRS distribution at each P_T_ were compared using t-tests. Thirdly, regression analysis described above was performed with each of the 10 P_T_ instead of PRS-PC1.

### 2.4. Exploratory gene-set enrichment analysis

To further examine if ASD common genetic variants most associated with lobe volumes converged on relevant biological pathways, exploratory gene-set enrichment analysis was carried out. Here, linear regression was performed for each SNP (Volume ∼ GA + PMA + TBV + Risk allele frequency + sex + ancestry PCs) to determine its effect on fronto-temporal and parieto-occipital lobe volumes.

Subsequently, only the SNPs nominally associated with the imaging phenotypes were considered for further analysis (p < 0.01). Here, three SNP subsets were examined: SNPs associated with only fronto-temporal lobe volume, SNPs associated with only parieto-occipital lobe volume, and SNPs associated with both volumes. To determine if elements in each SNPs subset converged on relevant biological pathways, genes containing those SNPs were probed for their functions. Here, a gene was selected if it contained the SNP of interest within its start and stop coordinates according to the human genome build 37 (https://ctg.cncr.nl/software/magma). Thus, for each SNPs subset, a corresponding gene list was created. Each gene list was then functionally tested against a curated database of 13 159 gene sets (pathways) obtained from MSigDB v.7.5.1 (curated canonical pathways from Reactome, KEGG, Wikipathways and Gene Ontology; https://www.gsea-msigdb.org/gsea/msigdb/). For each pathway, the hypergeometric test was used to determine the probability of randomly observing the set of genes in the gene list ^37^. This analysis was performed using GENE2FUNC on FUMA platform ^38^ with 19427 genes from human genome Build 37 as background genes. Pathways with adjusted Bonferroni p-value < 1.26 x 10^-6^ (i.e, 0.05/(13159 pathways *3 gene lists)) and containing at least 4 overlapping genes with the gene list were considered enriched. Finally, to ensure consistency and reproducibility of the results, two additional stability tests were carried out for each gene list. In the first test, we randomly sampled n SNPs (where n is the number of SNPs found in each SNPs subset), generated the corresponding gene list, and applied the hypergeometric method described above. This test was simulated 1000 times, and the most enriched pathway in each run was recorded. Gene sets specific to the phenotype of interest were those that were found across fewer than 5% of all random experiments. In the second test, we repeated the enrichment analysis of the gene lists with other similar bioinformatic tools, including DAVID v.2021 (http://david.abcc.ncifcrf.gov/) ^37,39^ and WebGestalt v. 2019 (http://www.webgestalt.org/#) ^40^ to confirm the FUMA findings.

## 3. Results

### 3.1 Higher ASD PRS associated with greater fronto-temporal volume but lower parieto-occipital volumes

We found statistically significant associations between frontal (β = 0.027, p_FDR_ = 0.04), parietal (β = −0.037, p_FDR_ = 0.04), fronto-temporal (β = 0.024, p_FDR_ = 0.01), and parieto-occipital lobe (β = −0.033, p_FDR_ = 0.01) volumes and PRS-PC1 (Figure 3). Here, higher PRS-PC1 was associated with higher frontal and fronto-temporal volumes, but lower parietal and parieto-occipital volumes. Performing stability tests by halving or comparing the top and bottom extremes of different PRS P_T_ distributions showed consistent direction of association in the reported brain regions (Supplementary Figure 2 and Supplementary Figure 3). Finally, consistent patterns of associations were also observed when considering all 10 PRS P_T_ values (Supplementary Figure 4).

**Figure 3.**
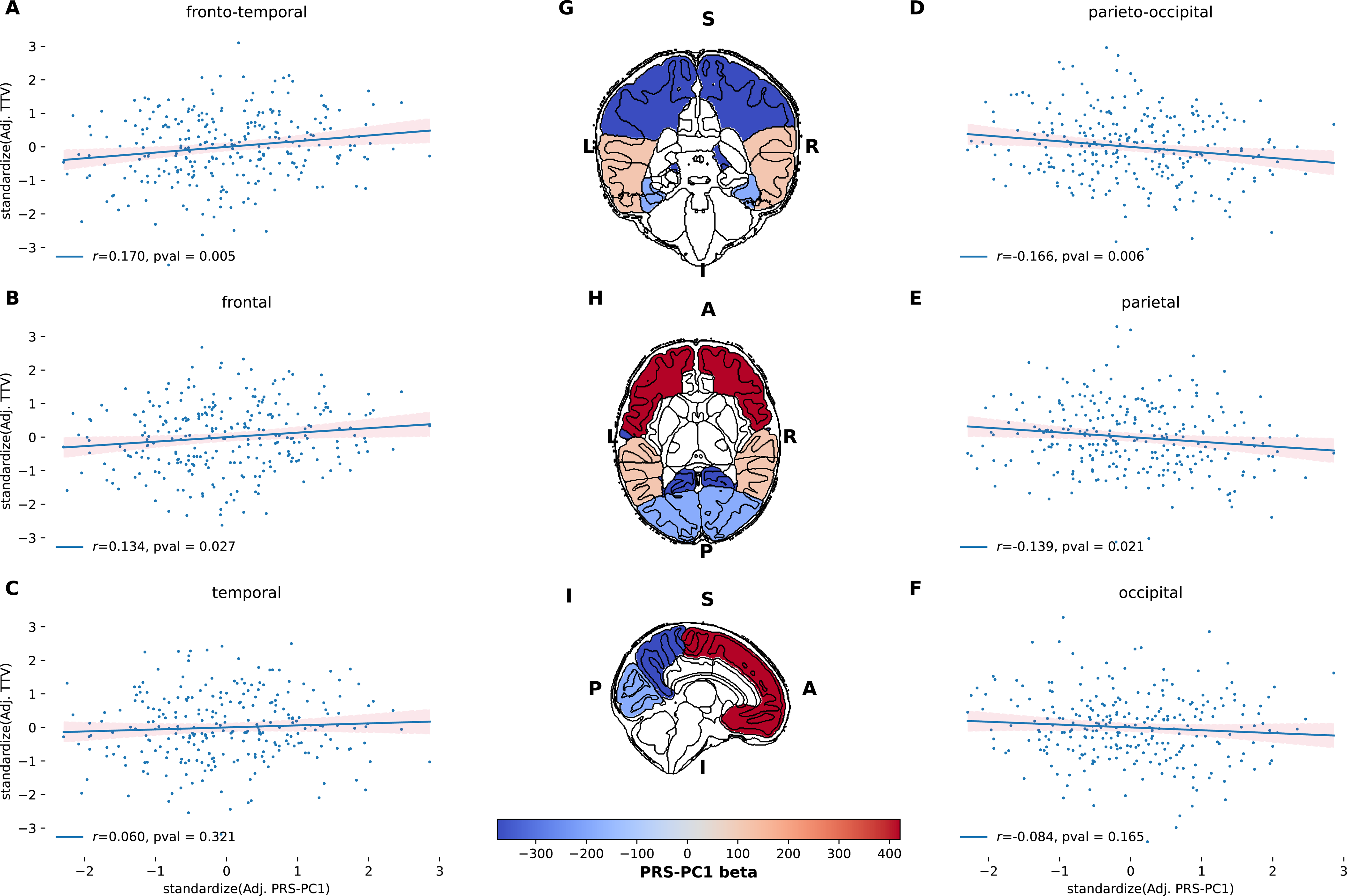
Association between PRS-PC1 and regional volumes. Left (A-C) and right (D-F) panels show scatterplots denoting correlation r and p-value between regional volumes and PRS-PC1 on y- and x-axis respectively. Values of regional volumes and PRS-PC1 have been standardized and adjusted for GA, PMA, sex, and TBV and first 3 ancestral PCs, respectively. Middle (G-I) show brain regions overlapped with values of PRS-PC1 beta coefficient in the linear regression model, where blue and red values indicate negative and positive direction of effect; S-superior, I-Inferior, A-anterior, P-posterior, L-left, R-right.

### 3.2 Enrichment in pathways related to neuron differentiation and synaptic membrane

From the 116 433 SNPs examined in this study, three unique SNPs subsets and their corresponding gene lists associated with fronto-temporal volume, parieto-occipital volume, or both were generated (Figure 4A). Overrepresentation analyses of the three gene lists identified enrichment in several gene-ontology biological processes and cellular components (Supplementary Table 1). Retaining only the significant pathways found in less than 5% of times across all random simulations revealed 2 gene sets, synaptic membrane (Gene Ontology (GO) cellular component, GO: 0097060; p=9.5×10^-7^) and neuron differentiation (GO biological process, GO: 0030182, p=7.6×10^-7^). Finally, gene sets enriched for similar functions were also found using both DAVID and WebGestalt tools (Supplementary Table 2 and 3).

**Figure 4.**
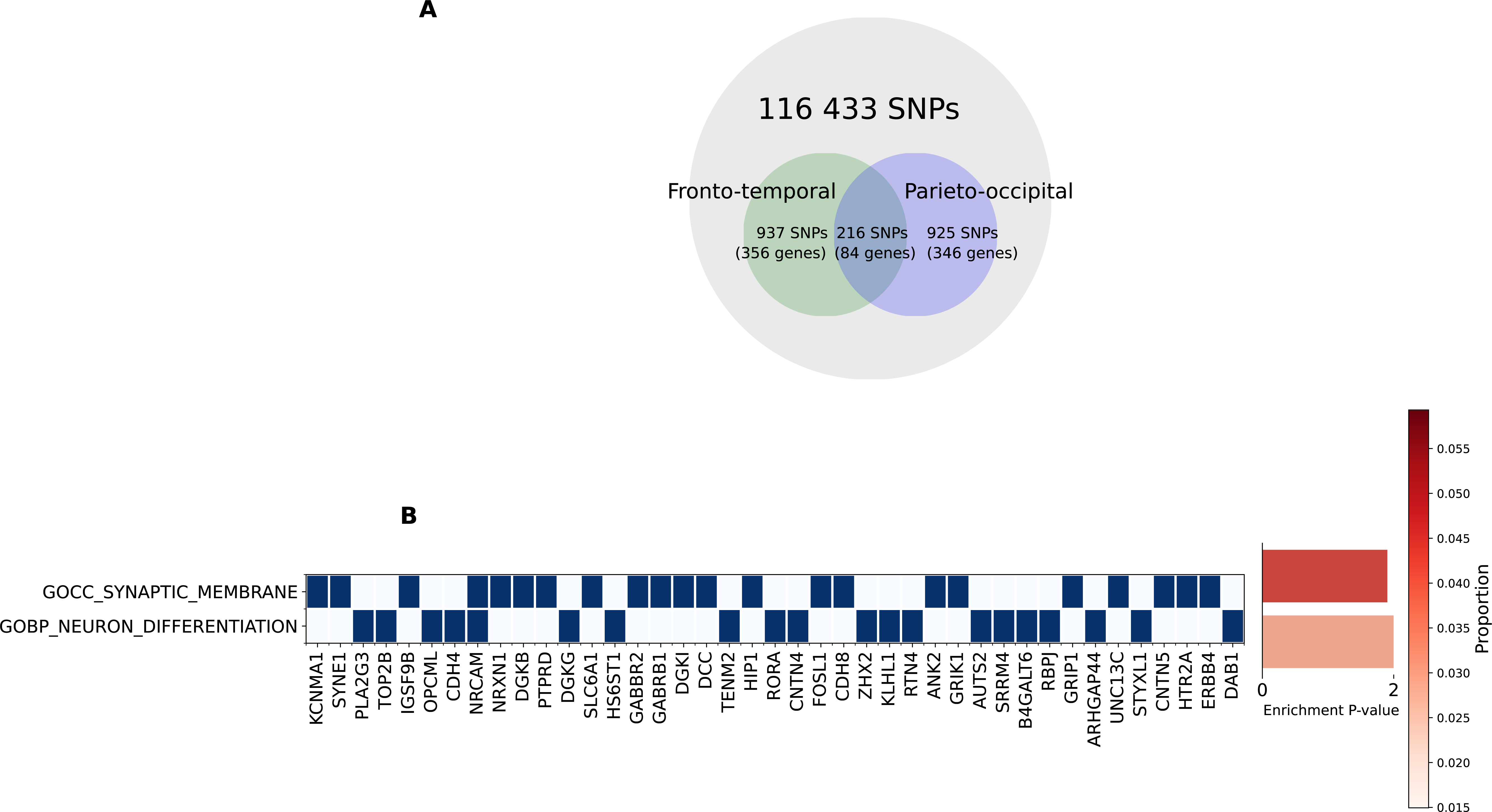
Gene set-enrichment analysis. A) Visualisation of the total SNPs examined in this study and number of SNPs and genes determined as associated with three phenotypes of interest B) Heatmap of genes involved in biological pathways enriched and specific (found in less than 5% across all random simulations) to the neuroimaging phenotypes of interest. Bar plot showing pathway enrichment p-value after Bonferroni correction and proportion of gene overlapped with the gene list. GOCC: Gene ontology cellular components. GOBP: Gene ontology biological pathways.

## 4. Discussion

We identified an association between ASD common genetic variants and regional brain lobe volumes in a cohort of term-born infants at birth. Here, we reported a positive association between fronto-temporal brain volume and ASD risk, but negative association of parieto-occipital brain volume and ASD risk. These results show measurable differences in brain volume associated with ASD risk prior to the onset of disease. In addition, exploratory gene set enrichment analysis of the ASD common variants most associated with the examined neuroimaging phenotypes suggested possible widespread disruption to normal early development processes.

### 4.1. Comparison with previous examination of ASD related structural variations

To our knowledge, this is the first study to examine the association of ASD common genetic variants with regional brain lobe volumes in a general population at birth. Previously, in two similar but larger cohorts of children between 3-14 and 9-12 years of age, ASD PRS has been significantly associated with cortical thickness of precentral and postcentral gyri ^2^ and nominally (i.e., did not survive multiple testing correction) associated with superior parietal gyrification ^41^. Taken together, these studies suggest that ASD common genetic variants may be involved in early structural brain changes.

Evidence of abnormal cortical surface area expansion has been observed in infants with a high-familial risk of ASD in the first year of life ^1^. In retrospective studies of ASD children, this surface area enlargement appeared to relate to cerebral brain volume increase in ASD children between 2 to 4 years old ^1,13^. In both examination of cortical surface area and cerebral lobe volumes, there appeared to be regional variability associated with the disease. This study provides additional evidence for regional brain volume differences associated with ASD PRS measurable as early as birth. While it is not yet known whether such brain structural changes persist in adolescence and adulthood ^42^, a recent longitudinal study of ASD cases versus controls between 2 and 13 years of age found that early cerebral enlargement was maintained until middle childhood with no evidence of normalisation or regression ^9^.

Greater frontal and fronto-temporal volume in association with greater ASD risk is in line with existing brain imaging studies ^43^. Evaluation of postmortem prefrontal cortices has suggested this may be driven by abnormally excess number of neurons in ASD children compared with that in controls ^44^. However, to our knowledge, no studies to date have identified reduced brain volume either in individuals with ASD or in association with ASD risk. While the magnitude of cerebral hyperplasia appears to vary across regions, with frontal and temporal lobes being more affected than the parietal and occipital lobes ^30^, our result of reduced parietal and occipital lobe volumes have not yet been reported. Given that previous exploration of ASD PRS has focused on total brain volume and global grey and white matter ^41,45^, we could not draw comparison with our current result. Interestingly, recent transcriptomic analysis of postmortem ASD brain tissues across four cortical lobes similarly found evidence of variability in expression of risk genes along the antero-posterior axis ^17^. Taken together, this suggests pathophysiology of the disease may involve disruption to normal neuro-patterning process. Concurrently, the current study reported a consistent pattern of opposite directions of effect related to ASD common variants between fronto-temporal and parieto-occipital regions.

### 4.2 Clinical relevance of neuroimaging observations with pathophysiology of ASD

While the pathophysiology of ASD likely begins prenatally, the evidence suggests that subtle ASD-traits emerge during the latter part of the first and second years in high-risk infants who later develop the disease ^46^. For instance, at 6 months, high-risk infants exhibit differences in gross motor and visual reception skills ^47^. Interestingly, recent studies also found significant association between ASD PRS and motor and language scores in general infant populations at 18 months^48,49^, raising the possibility of partial contributions by ASD common variants on related ASD-traits. However, the current consensus is that ASD-defining features likely become pronounced only after the second year and consolidate around the fourth year, which co-occur with the regional brain volume overgrowth ^46^. Indeed, disruptions to the regions examined in this study have been implicated in reduced social and language functions and repetitive or stereotyped behaviors of ASD ^50^. Nevertheless, the direction and effect of association between regional brain lobe volumes, ASD symptoms and ASD common variants remain largely unclear and future investigation into this interplay in early life may provide important insights into the pathophysiology of the disease.

### 4.3 Exploratory gene set enrichment analysis

By selecting the ASD common variants that were associated with fronto-temporal and parieto-occipital volumes, we found enriched neurodevelopmental processes. However, this is expected since ASD common variants are likely found in genes primarily involved in corticogenesis and with highest expression during the prenatal period ^15^. In addition, we have previously identified regional brain volume variations associated with schizophrenia PRS in the same cohort of infants ^51^. Together, these preliminary results support the notion that many psychiatric disorders may have neurodevelopmental origin and may be associated with brain structural variations before the average age of the disease diagnosis ^20^.

Dysregulation of multiple aspects of synaptic function and neuron development have been implicated in the disease. For instance, combinations of abnormal synaptogenesis (e.g. due to mutations in core regulators of synaptic architectures) and synaptic pruning (e.g., defects in neuronal autophagy) have been proposed to lead to increased density of dendritic spines in ASD patients ^52^. Similarly, abnormalities in the control of neuronal cell differentiation may give rise to accelerated neuronal cell growth observed in ASD ^53^.

### 4.4. Limitations

The main limitation of our study is the small sample size and simplicity of PRS. Therefore, findings presented here must be considered preliminary, and confirmation in independent samples is important. Here, only term-born infants of European ancestry were considered. This was firstly due to the known impacts of prematurity on the brain structures and secondly due to exclusive inclusion of Danish population in the discovery GWAS sample. In line with the latest schizophrenia GWAS ^54^, future inclusion of different ancestral groups in the ASD GWAS may improve generalisability of the PRS. Finally, while overrepresentation analyses must be considered exploratory, as the analyses are biased toward larger well-studied processes ^37^, they may still provide relevant targets for future investigation.

## 5. Conclusion

In summary, our study reports positive associations between the ASD PRS with the frontal and fronto-temporal volumes, but negative with the parietal and parieto-occipital volumes in a cohort of term neonates. Whilst preliminary, the result contributes to the examination of the pathophysiology of ASD and its early emergence.

## Data availability

The MRI data used (dHCP third release) is freely available: https://biomedia.github.io/dHCP-release-notes/. The described statistical analysis can be recreated using the IPython notebook https://github.com/lehai-ml/dHCP_genetics/blob/main/notebook_results/asd/Supplementary_notebook.ipynb. Scripts used for visualising results in this study is available here: https://github.com/lehai-ml/nimagen.

## Acknowledgements

Data were provided by the developing Human Connectome Project, KCL-Imperial-Oxford Consortium and the work was funded by ERC grant agreement no. 319456, the Wellcome EPSRC Centre for Medical Engineering at Kings College London (WT 203148/Z/16/Z) and by the National Institute for Health Research (NIHR) Biomedical Research Centre based at Guy’s and St Thomas’ NHS Foundation Trust and King’s College London. The views expressed are those of the authors and not necessarily those of the NHS, the National Institute for Health Research or the Department of Health. The funders had no role in the design and conduct of the study; collection, management, analysis, and interpretation of the data; preparation, review, or approval of the manuscript; and decision to submit the manuscript for publication. We are grateful to the families who generously supported this trial.

HL is supported by the UK Medical Research Council (MR/N013700/1) and King’s College London member of the MRC Doctoral Training Partnership in Biomedical Sciences.

HC is an academic clinical lecturer in Clinical Genetics at Kings College London and is supported by the NIHR. LCG received support from the Comunidad de Madrid-Spain Support for R&D Projects [BGP18/00178].

The authors acknowledge use of the research computing facility at King’s College London, Rosalind (https://rosalind.kcl.ac.uk).

