## Supplementary Information for "Autism spectrum disorder common variants associated with regional lobe volume variations at birth: cross-sectional study in 273 European term neonates in developing Human Connectome Project"

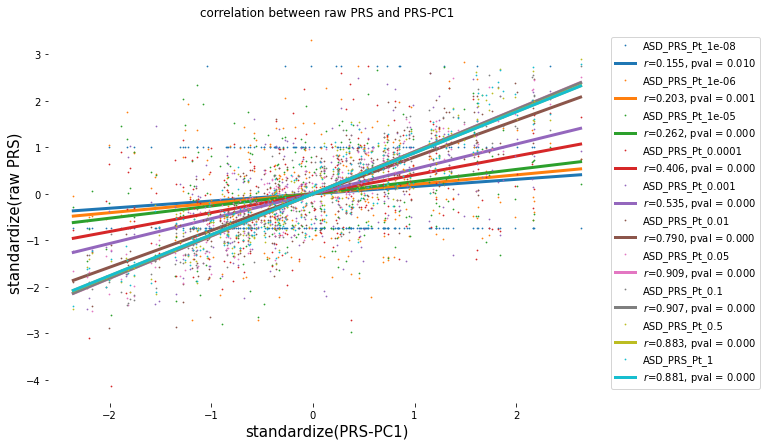
Supplementary Figure 1. Correlation between PRS-PC1 and raw PRS scores. Y-axis – standardised values of all 10 P_T_. X- axis - standardised values of the PRS-PC1. Legend shows *r* - correlation and *p-value* between PRS-PC1 and each respective PRS P_T_.

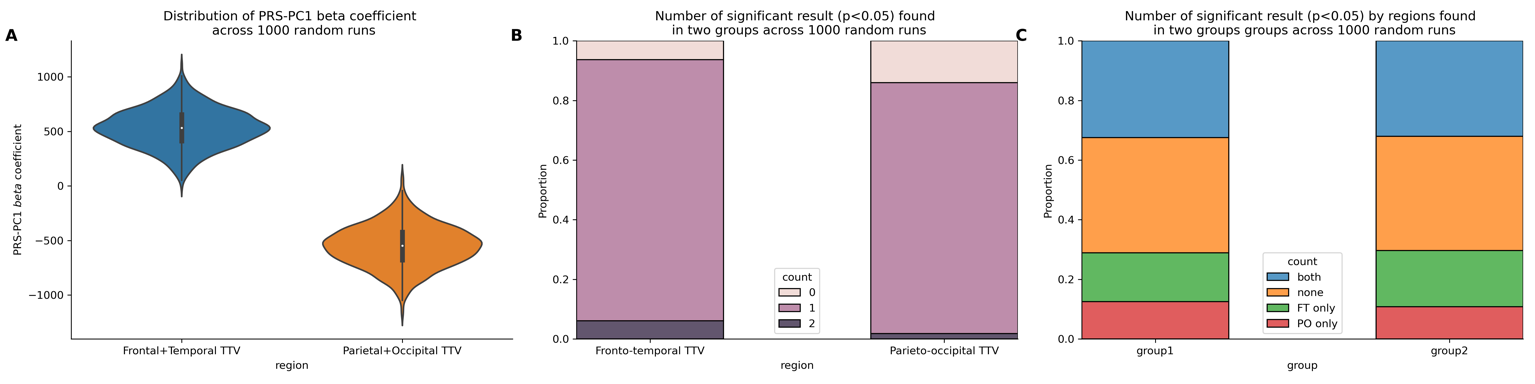
Supplementary Figure 2. Visualisation of the 1000 random regression analyses. Here, the total data set is randomly divided into 2 datasets. PCA is applied to each of the datasets separately to yield PRS-PC1. Then each dataset is fitted in linear regression, TTV ~ PRS-PC1 + Ancestry PCs + GA + PMA + TBV + sex. A) Violin plot of the distribution of the PRS beta coefficients associated with either fronto-temporal or parieto-temporal TTV across 2 datasets x 1000 regression = 2000 runs. B) Bar plot showing the proportion of number of significant results (p<0.05) associated with either Fronto-temporal TTV or Parieto-occipital TTV found in either 1 or both data sets and none across 1000 random regression simulations. Here, of 1000 random run, in 937 and 860 runs we found significant association between PRS-PC1 and fronto-temporal TTV and Parieto-occipital TTVs, respectively, in at least one dataset. Of which, 61 and 18 runs found significant association in both datasets. C) Number of significant associations found in each dataset (group). In dataset 1- of the 1000 random simulation, we found 325 runs where association (p<0.05) was found between PRS-PC1 and fronto-temporal and parieto-occipital TTVs, 164- with only fronto-temporal, 125 – only with parieto-occipital and 386 - no association with either phenotype. In dataset 2- of the 1000 random simulation, we found 320 runs –association in both phenotypes, 189 – fronto-temporal only, 108 – parieto-occipital only, and 383 – neither.

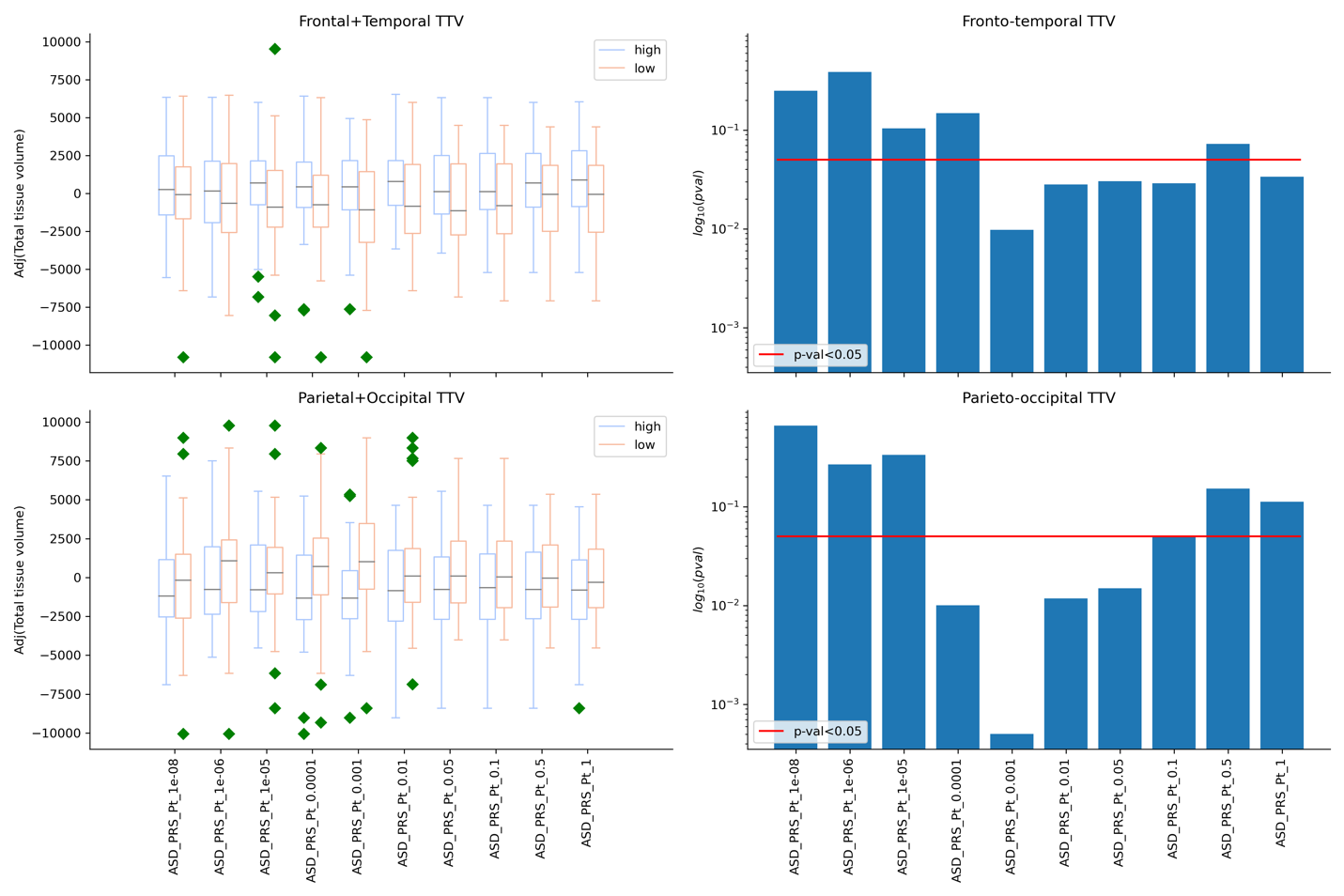

Supplementary Figure 3. Differences in TTVs of the top and bottom 20% PRS. Left panels – Box plot visualizing differences in fronto-temporal (top) and parieto-occipital (bottom) TTVs between high risk (top 20% of the adjusted PRS score) and low risk (bottom 20% of the adjusted PRS score) across multiple PRS P_T_. Here, the Right panels – Bar plot visualizing the p-value in t-test for 2 independent samples comparing the high (top 20%) vs. low (bottom 20%) risk across all 10 P_T_. Red line denotes the p-value < 0.05. Y-axis log_10_(t-test p-value).

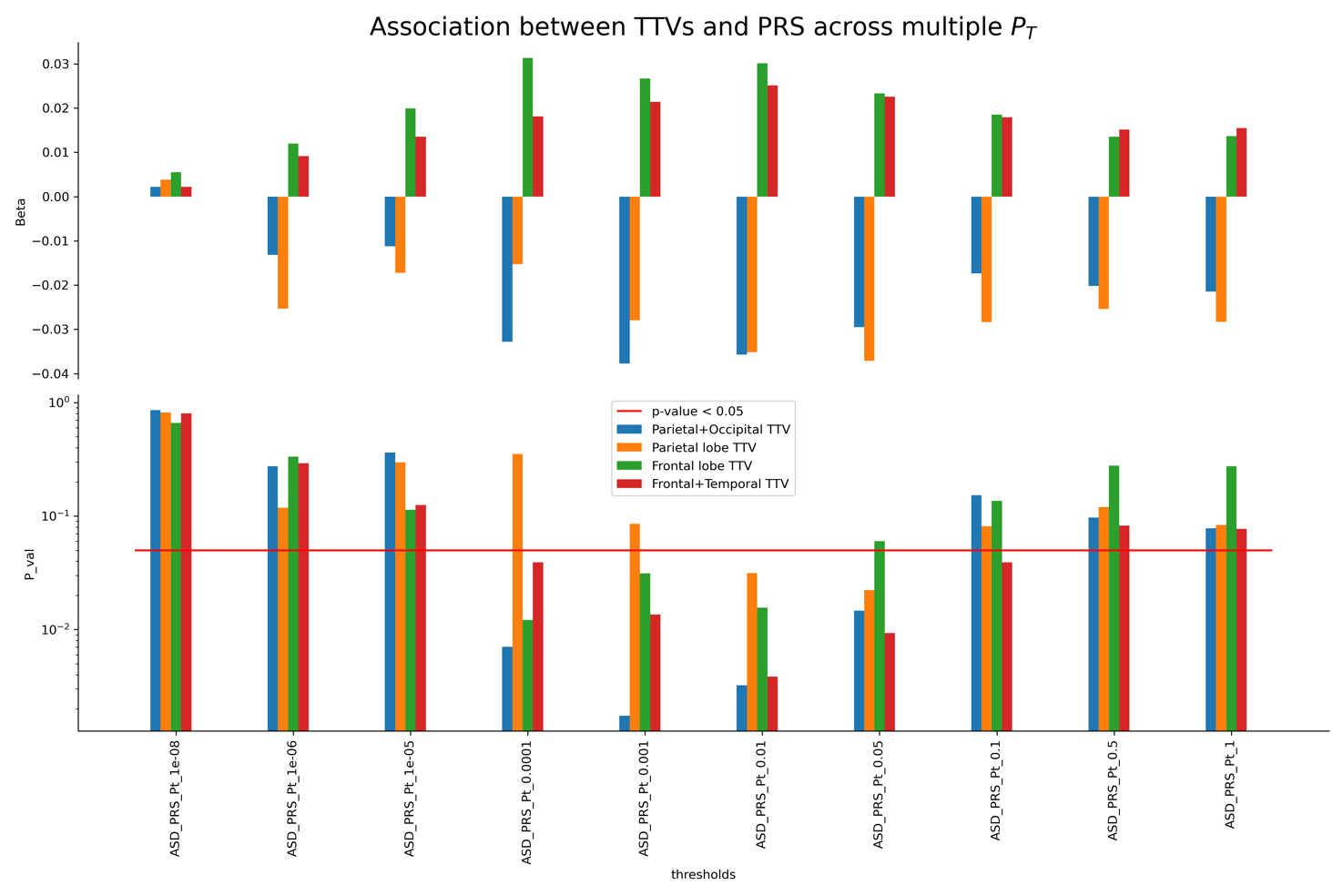
Supplementary Figure 4. Association between 4 TTVs (frontal, parietal, fronto-temporal, parieto-occipital lobes) and PRS across all 10 P_T_. Linear regression was fitted for each of the 10 P_T_, such that TTV ~ PRS + Ancestry PCs + GA + PMA + sex + TBV. Top plot – standardised PRS beta coefficient in the linear regression model. Bottom plot – log_10_(p-value). Red line: Nominal significance – p-value < 0.05.
